# Serologic SARS-CoV-2 testing in healthcare workers with positive RT-PCR test or Covid-19 related symptoms

**DOI:** 10.1101/2020.10.25.20219113

**Authors:** Giovanni Visci, Vittorio Lodi, Roberta Bonfiglioli, Tiziana Lazzarotto, Francesco S. Violante, Paolo Boffetta

## Abstract

**Background:** Limited information is available on prevalence and determinants of serologic response to SARS-CoV-2 infection among healthcare workers (HCWs).

**Methods:** We analyzed the results of serologic testing with chemiluminescence immunoassay analyzer (CLIA), lateral flow immunoassay (LFIA) and enzyme-linked immunosorbent assay (ELISA) test among 544 HCWs with at least one positive RT-PCR test and 157 HCWs with Covid-19 related symptoms without a positive RT-PCR test from public hospitals in Bologna, Northern Italy. Tests were performed between March and August 2020. We fitted multivariate logistic regression models to identify determinants of positive serology.

**Results:** The sensitivity of SARS-CoV-2 was 75.2% (LFIA) and 90.6% (CLIA). No differences in seropositivity were observed by sex, while older HCWs had higher positivity than other groups, and nurses had higher positivity compared to physicians, but not other HCWs. An estimated 73.4% of HCWs with Covid-19 symptoms without RT-PCR test were not infected with SARS-CoV-2.

**Conclusions:** Our study provides the best available data on sensitivity of serologic tests and on determinants of serologic response among HCWs positive for SARS-CoV-2, and provide evidence on the low specificity of Covid-19 related symptoms to identify infected HCWs.

**Summary:** The sensitivity of SARS-CoV-2 lateral flow immunoassay serology in healthcare workers (HCWs) was 75.2%. Older HCWs and nurses had higher positivity than other groups. An estimated 73.4% of HCWs with Covid-19 symptoms without RT-PCR test were not infected with SARS-CoV-2.

## Introduction

Infection with the virus, Severe Acute Respiratory Syndrome Coronavirus-2 (SARS-CoV-2), which may cause Coronavirus Disease 2019 (Covid-19), spread extensively following its identification in December, 2019, becoming a global pandemic by March, 2020.

Healthcare workers (HCWs) are considered a high-risk population for the acquisition of SARS-CoV-2 infection, due to the probability and potentially high level of exposure associated with clinical care of positive cases and infected coworkers. Use of personal protective equipment (PPE) and stringent infection prevention and control measures aim to mitigate the risk and minimize both workplace-related infection of HCWs and onward transmission [1].

During the initial period, when the incidence of new cases was high, but reverse transcriptase polymerase chain reaction (RT-PCR) testing was not yet widely available, a large proportion of symptomatic Italian HCWs were not tested. Thus, SARS-CoV-2 prevalence in this population remains poorly defined. Few studies have reported either current or past infection status using both RT-PCR and serological testing [2, 3]. One meta-analysis has summarized the results of studies on serologic response in patients with Covid-19 [4]; however, little is known about the seroconversion rate following asymptomatic infection in health care personnel. A recent Cochrane Review assessed the diagnostic accuracy of antibody tests to determine current or previous SARS-CoV-2 infection and their use in seroprevalence surveys. The sensitivity of antibody tests was found too low in the first week since symptom onset to have a primary role for the diagnosis of COVID-19. However, antibody tests may have a role complementing other testing in individuals presenting later, when RT-PCR tests are negative, or are not done and for detecting previous SARS-CoV-2 infection if used 15 or more days after the onset of symptoms. Most studies included hospitalized patients, little is known among asymptomatic cases [5].

In the present study we aimed to analyze the prevalence of positive serology testing following positive RT-PCR or the appearance of symptoms suggestive of Covid-19 among high-risk HCWs employed in public hospitals of Bologna, Northern Italy, an area at high incidence of infection with SARS-CoV-2 and mortality from Covid-19, and to compare the sensitivity of different types of serological tests, including chemiluminescence immunoassay analyzer (CLIA), lateral flow immunoassay (LFIA) and enzyme-linked immunosorbent assay (ELISA).

## Methods

The study consisted of a retrospective analysis of RT-PCR results and serology data collected among HCWs employed in a large university hospital, or in a specialized orthopedic hospital and other public hospitals in Bologna, who were included in a surveillance program managed by the Occupational Health Unit of the university hospital.

HCWs who experienced a close contact with a confirmed case of Covid-19 (whether a coworker or a patient) and/or exhibited symptoms compatible with Covid-19 (either two major symptoms, including cough, sore throat, fever, myalgia, asthenia, anosmia, ageusia, and dyspnea, or one major and two minor symptoms, including rhinorrhea, chills, arthralgia, diarrhea, conjunctivitis, and vesicular erythema) were tested for SARS-CoV-2 infection and were included in a surveillance program, that included telephone contacts for symptoms monitoring and, where required, the prescription of medications. Nasopharyngeal swab/oropharyngeal swab samples were analyzed by RT-PCR according to the guidelines proposed by the World Health Organization [6]. An additional group of healthcare workers who developed symptoms related to Covid-19, but did not have a positive RT-PCR result, was included in the surveillance program (based on clinical diagnostic criteria). Some of the HCWs in the latter group were tested by RT-PCR at the end of the 14-day surveillance period, with a negative result.

The two groups underwent antibody test using one or more out of three different methods, including CLIA, LFIA and ELISA. The protocol included first an CLIA or a LFIA test, followed, in case of positive result, by a confirmatory ELISA test. Due to changes in the diagnostic methods over time, a subgroup of HCWs who tested negative at LFIA or CLIA, or not having been tested, underwent an ELISA test for detecting antibodies to SARS-CoV-2. Conversely, some HCWs who tested positive at LFIA or CLIA did not undergo an ELISA test.

The tests used were the SARS-CoV-2 IgM and IgG CLIA kits (Shenzhen YHLO Biotech, Shenzhen, China), with cutpoint for positivity at 10 AU/ml for IgG and IgM combined, the nCOVID-19 IgG and IgM POCT (Technogenetics, Milan, Italy) LFIA, and the ELISA (DiaPro, Milan, Italy) test, the latter with cutpoint for positivity for IgG at 3 signal/threshold ratio.

RT-PCR testing and identification of symptomatic HCWs were performed between 6 March 2020 and 6 July 2020, while serological tests for detecting antibodies to SARS-CoV-2 were performed between 2 April 2020 and 4 August 2020.

The outcome of the present analysis was the presence of at least one positive result for anti-SARS-CoV-2 (IgG for LFIA and ELISA; IgG/IgM for CLIA, which does not distinguish between the different classes of Ig). Each HCWs entered in the study on the day of the first positive RT-PCR test or, for those without such test, the day when Covid-19-related symptoms were first reported. For each subject, we ignored the results of antibody tests conducted less than 7 days following the enrollment. For each type of test, we assessed whether the HCWs with valid results had at least one positive results or only negative results.

We fitted multivariable logistic regression models to the data to estimate odds ratios (OR) of a positive serology result for each type of test, together with 95% confidence intervals (CI). All regression models included sex, age (categorized as 20-34, 35-44, 45-54, and 55 and over), job title (physician, nurse, health worker, and other jobs), date of first positive RT-PCR test (categorized as 6 March – 18 March, 19 March – 29 March, 30 March – 19 April, and 20 April – 6 July; a comparable variable was not available for HCWs without RT-PCR test because information on symptom onset was self-reported, and therefore subject to misclassification), and the institution (university hospital, orthopedic hospital, other public hospitals) as potential confounders.

We also estimated the proportion *p* of HCWs with Covid-19 related symptoms, who were not infected with SARS-CoV-2, using the formula

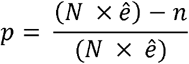

where *N* is the total number of HCWs with Covid-19 related symptoms who were tested with LFIA, *ê* is the estimated sensitivity of the LFIA test (based on the results among HCWs with either a positive RT-PCR test or a positive ELISA result), and *n* is the number of HCWs with Covid-19 related symptoms who tested positive.

## Results

A total of 701 HCWs were included in the study, of whom 544 (77.6%) had at least one positive RT-PCR test and 157 (22.4%) had Covid-19 related symptoms without a RT-PCR test. Among the 701 HCWs, 74 had at least one ECLIA test, 660 had at least one LFIA test, and 458 had at least one ELISA test. Among HCWs who underwent CLIA testing, 58 (74.4%; 95% confidence interval [CI], 69.0% - 87.8%) had at least one positive result. Among 660 HCWs with at least one LFIA test, 403 (61.1%; 95% CI 57.3% - 64.8%) had at least one positive results; this proportion was 91.5% (95% CI, 88.9% - 94.0%; 419 positive results out of 458 HCWs) in those with at least one ELISA test.

The estimate of the sensitivity of LFIA test was 75.2% (95% CI 71.3%-78.8%, based of 403 positive results among 536 HCWs with either a positive RT-PCR test or a positive ELISA result); the estimate of the specificity was 100% (95% CI 85.2%-100%, based on 23 negative results among 23 HCWs with no RT-PCR test and only negative ELISA results). Corresponding estimates for CLIA test were 90.6% (95% CI 80.7%-96.5%; 58/64 HCWs) and 100% (95% CI 47.8%-100%; 5/5 HCWs).

The proportions of positive results for the three types of serologic test according to the criterion for inclusion in the study and selected characteristics are reported in Table 1, and the results of the multivariable logistic regression for LFIA and ELISA test among HCWs with positive RT-PCR test are reported in Table 2 (results of the corresponding analysis of results of CLIA tests were hampered by small numbers, resulting in unstable estimates, and are not reported in detail). The overall proportion of positive results was higher for all three types of test among HCWs with a positive RT-PCR test (86.9% for CLIA, 73.7% for LFIA and 96.3% for ELISA) than among HCWs with Covid-19 related symptoms (38.5%, 20.0% and 57.1%, respectively, test of the difference between two groups of HCWs, p = 0.001 for CLIA and p < 0.00001 for LFIA and ELISA).

**Table 1.** Results of LFIA and ELISA test among HCWs with positive RT-PCR test.

| LFIA test | ELISA test |  |  | Total |
| --- | --- | --- | --- | --- |
|  | Positive | Negative | Not performed |  |
| Positive | 399 | 3 | 1 | 403 |
| Negative | 5 | 33 | 219 | 257 |
| Not performed | 15 | 3 |  | 18 |
| Total | 419 | 39 | 220 | 678 |

LFIA, lateral flow immunoassay ELISA, enzyme-linked immunosorbent assay

**Table 2.** Results of serologic test by selected characteristics of HCWs.

|  |  | HCWs with positive RT-PCR<br>(positive/negative) |  |  | HCWs with Covid-19 related<br>symptoms (positive/negative) |  |  |
| --- | --- | --- | --- | --- | --- | --- | --- |
|  |  | CLIA<br>(N=61) | LFIA IgG<br>(N=505) | ELISA IgG<br>(N=402) | CLIA<br>(N=13) | LFIA IgG<br>(N=155) | ELISA IgG<br>(N=56) |
| Sex | Female | 42/5 | 253/93 | 266/9 | 2/5 | 22/80 | 22/17 |
|  | Male | 11/3 | 119/40 | 121/6 | 3/3 | 9/44 | 10/7 |
| Age | 20-34 | 16/5 | 91/52 | 96/4 | 3/1 | 9/32 | 10/8 |
|  | 35-44 | 12/1 | 80/31 | 84/3 | 0/2 | 7/46 | 7/7 |
|  | 45-54 | 19/1 | 114/32 | 120/5 | 0/4 | 10/28 | 10/7 |
|  | 55+ | 6/1 | 87/18 | 87/3 | 2/1 | 5/18 | 5/2 |
| Institution | Other public hospitals | 24/4 | 209/55 | 211/8 | - | - | - |
|  | University hospital | 29/4 | 111/58 | 124/7 | 5/7 | 19/90 | 20/23 |
|  | Orthopedic hospital | 0/0 | 52/20 | 52/0 | 0/1 | 12/34 | 12/1 |
| Job title | Nurse | 27/6 | 173/51 | 180/5 | 2/4 | 13/57 | 14/13 |
|  | Physician | 9/1 | 88/44 | 90/6 | 3/2 | 4/26 | 4/5 |
|  | Health worker | 9/0 | 64/19 | 67/3 | 0/1 | 7/22 | 7/4 |
|  | Other | 8/1 | 47/19 | 50/1 | 0/1 | 7/19 | 7/2 |
| Date of RT-PCR test | 6 March-18 March | 5/1 | 92/59 | 92/8 | - | - | - |
|  | 19 March-29 March | 10/2 | 129/27 | 130/2 | - | - | - |
|  | 30 March-19 April | 21/1 | 115/26 | 124/3 | - | - | - |
|  | 20 April-6 July | 17/4 | 36/21 | 41/2 | - | - | - |
| Total |  | 53/8 | 372/133 | 387/15 | 5/8 | 31/124 | 32/24 |
| Proportion positive | % | 86.9 | 73.7 | 96.3 | 38.5 | 20.0 | 57.1 |
|  | 95% CI | 78.4-95.4 | 69.8-77.5 | 94.4-98.1 | 12.0-64.9 | 13.7-26.3 | 44.2-70.1 |

ECLIA, electro-chemiluminescence immunoassay
LFIA, lateral flow immunoassay
ELISA, enzyme-linked immunosorbent assay
CI, confidence interval

**Table 3.** Determinants of positive serology test – Results of multivariable logistic regression restricted to HCWs with positive RT-PCR test.

|  |  | LFIA IgG |  | ELISA IgG |  |
| --- | --- | --- | --- | --- | --- |
|  |  | OR | 95% CI | OR | 95% CI |
| Sex | Female | 1 | Ref. | 1 | Ref. |
|  | Mamle | 1.39 | 0.86-2.24 | 0.85 | 0.27-2.62 |
| Age | 20-34 | 1 | Ref. | 1 | Ref. |
|  | 35-44 | 1.31 | 0.74-2.32 | 1.19 | 0.23-6.22 |
|  | 45-54 | 1.92 | 1.09-4.00 | 0.93 | 0.22-3.93 |
|  | 55+ | 3.35 | 1.73-6.49 | 1.71 | 0.34-8.50 |
| Institution | Other public hospitals | 1 | Ref. | 1 | Ref. |
|  | University hospital | 0.46 | 0.29-0.75 | 0.66 | 0.22-2.01 |
|  | Orthopedic hospital | 0.76 | 0.40-1.47 | ∞ | - |
| Job title | Nurse | 1 | Ref. | 1 | Ref. |
|  | Physician | 0.57 | 0.33-0.96 | 0.37 | 0.10-1.39 |
|  | Health worker | 0.81 | 0.43-1.53 | 0.47 | 0.10-2.18 |
|  | Other | 0.62 | 0.31-1.25 | 1.01 | 0.11-9.25 |
| Date of RT-PCR test | 6 March-18 March | 1 | Ref. | 1 | Ref. |
|  | 19 March-29 March | 3.05 | 1.76-5.30 | 6.59 | 1.32-32.8 |
|  | 30 March-19 April | 3.64 | 2.03-6.53 | 5.22 | 1.26-21.7 |
|  | 20 April-6 July | 1.19 | 0.60-2.34 | 2.17 | 0.41-11.4 |

LFIA, lateral flow immunoassay
ELISA, enzyme-linked immunosorbent assay
OR, odds ratio adjusted for the factors in the table
CI, confidence interval;
Ref., reference category

No differences in the proportion of positive results were observed by sex, while older HCWs tended to have a higher proportion of positive antibody results compared to younger subjects – although the difference was not statistically significant for results of ELISA results. Compared to nurses, physicians had a lower proportion of positive results (not significantly so for ELISA test), while no differences were detected for other HCWs. The proportion of HCWs with a positive antibody test was highest among those tested with RT-PCR between 8 March and 19 April, and decreased among those tested later.

Based on the results reported in Table 2, and sensitivity of LFIA test equal to 75.2%, we estimated that 73.4% of HCWs with Covid-19 related symptoms, who were not tested with RT-PCR, were not infected with SARS-CoV-2.

## Discussion

In this population of HCWs with positive RT-PCR test for SARS-CoV-2 infection from three institutions in Bologna, Italy, the sensitivity of serologic LFIA test was 75.2% and that of CLIA test, based on smaller number of tests, was 90.6%. It is not possible to separate negative results due to lack of seroconversion from those due to false negative tests.. In the group of HCWs who exhibited Covid-19 related symptoms but were not tested with RT-PCR, the proportion of positive results for these two tests were 20.0%, and 38.5%, respectively. The lower proportion of positive serologic tests among HCWs with Covid-19 related symptoms compared to that of HCWs with RT-PCR test is likely due to lack of infection with SARS-CoV-2.The estimated specificity of LFIA and CLIA tests, based on small samples of HCWs likely negative for SARS-CoV-2 infection, was 100% for both tests.

Because of the design of the surveillance program, it is not possible to estimate the sensitivity and specificity of ELISA test. However, since most of the HCWs tested with ELISA had a positive LFIA result, it is likely that the proportion of positive ELISA results in our study overestimates the true sensitivity of the test.

In a systematic review and meta-analysis of studies of serologic results in patients with Covid-19, Lisboa Bastos and colleagues calculated a pooled sensitivity of 84.3% (95% CI 75.6%-90.9%) for ELISA tests measuring IgG or IgM, 66.0% (95% CI 49.3%-79.3%) for LFIA tests, and 97.8% (95% CI 46.2%-100%) for CLIA tests [4]. Pooled specificity ranged from 96.6% to 99.7%. To our knowledge, four studies reported results of serological tested among RT-PCR-positive HCWs. In a series from Italy, Lahmer and colleagues reported that 80% of RT-PCR positive HCWs were positive for ELISA IgG after an interval of 14 days; this proportion increased to 100% after an interval of 20 days (the exact number of HCWs was not reported but was lower than 58) [7]. In a study from Germany, 21/27 (78%) HCWs tested positive at RT-PCR, displayed also an IgG-positive ELISA test [8]. Another study found among 126 RT-PCR positive HCWs from Belgium, a proportion of positive CLIA test of 86.5% [9]. Finally, in a study of 340 RT-PCR positive HCWs from Denmark, a total of 334 subjects were tested positive for total ELISA Ig [10]. Our results, based on either LFIA or ECLIA, testing the largest group of RT-PCR HCWs studied so far, are comparable to those of previous studies.

The regression analysis, based on LFIA, offers the most informative figures with respect to determinants of positive anti-SARS-CoV-2 antibodies; the fact that they are consistent with the results of the comparable analysis ELISA, provides additional support. To our knowledge, these are the first results reported in the scientific literature on determinants of serologic response among HCWs infected by SARS-CoV-2, and are in line with similar analysis conducted among HCWs regardless of infection status [8,10-12].

It is well established that the main clinical manifestations of Covid-19 are fever (90% or more), cough (around 75%), and dyspnea (up to 50%) [13,14]. In addition, as demonstrated in a recent review by Meng and colleagues, olfactory dysfunction is a characteristic sign of Coid-19 patients, which can occur independently from other symptoms, even if its pathogenesis is not well understood [15]. In the scientific literature, on the other hand, there are few studies that correlate the presence of suspected symptoms for Covid-19 with the probability of having contracted this infection. In a model-based study, Sun and colleagues [16] found that elevated body temperature was the strongest predictor of Covid-19, in a study of HCWs, Lan and colleagues [17] found that fever, anosmia/ageusia, and myalgia were the strongest predictors of SARS-CoV-2 infection, while isolated sore throat and nasal congestion were negatively associated with the infection. In our study approximately 73% of HCWs with Covid-19 related symptoms, who were not tested with RT-PCR, were not infected with SARS-CoV-2. This is not surprising given the relatively low prevalence of SARS-Cov-2 infection, both in the general population and even among HCWs, a group at increased risk of infection. In a separate analysis we calculated at 5.8% the prevalence of RT-PCR positive results among the same group of workers infected up to early June 2020 [18]. The corresponding cumulative incidence of infection in the population of the province of Bologna, measured on 6 July 2020, was 0.52% [19]. Our work shows the difficulty in structuring a clinical and laboratory evaluation approach aimed at identifying suspected cases of Covid-19 only on clinical basis.

Our study suffers from several limitations. Data were collected during medical surveillance established in response to the first wave of SARS-CoV-2 pandemic during the spring of 2020, rather than within an *ad hoc* designed study. Furthermore, the protocol established for the surveillance of HCWs prevented us from analyzing the sensitivity of the ELISA test, that was mostly performed following a positive result based on LFIA. An additional limitation is that the current reference standard for SARS-CoV-2 infection confirmation, has a reported proportion of false negatives as low as 2% and as high as 37% according to the time of examination with respect to symptoms, if any [20].

In addition to the large sample size, strengths of the study include the fact that the same tests were consistently used for all HCWs, and analyzed in a single laboratory. Furthermore, data on potential determinants of serological results were collected before the results of the tests were known, suggesting that any misclassification was likely to be non-differential and lead to an underestimate of the associations.

Testing plays a vital role in the clinical management of Covid–19; in particular among high-risk groups such as HCWs, and the availability of serologic assays to detect antibodies against SARS-CoV-2 provide us additional tools in response to the Covid-19 pandemic. Our results add substantially to the available data on sensitivity of serologic LFIA and CLIA tests and on determinants of serologic response among HCWs confirmed SARS-CoV-2 cases and provide evidence on the low specificity of using Covid-19 related symptoms to identify infected HCWs.

## Data Availability

All data are extracted from a cross between Bologna metropolitan dashboard and health surveillance data extractions.
We may decide to deposit the dataset on Dryad

## Acknowledgments

The authors have no conflicts of interest.

Carlotta Zunarelli, Giulia Di Felice, Carmine Mastrippolito helped in data collection and management.

## References

1. Public Health England. COVID-19: infection prevention and control guidance. Updated 16 September 2020. [https://www.gov.uk/government/publications/wuhan-novel-coronavirus-infection-prevention-and-control/updates-to-the-infection-prevention-and-control-guidance-for-covid-19]

2. Treibel TA, Manisty C, Burton M, et al. COVID-19: PCR screening of asymptomatic health-care workers at London hospital. Lancet 2020;395:1608e10.

3. Korth J, Wilde B, Dolff S, et al. SARS-CoV-2-specific antibody detection in healthcare workers in Germany with direct contact to COVID-19 patients.J Clin Virol Off Publ Pan Am Soc Clin Virol 2020;128:104437.

4. Lisboa Bastos M, Tavaziva G, Abidi SK, et al. Diagnostic accuracy of serological tests for covid-19: systematic review and meta-analysis. BMJ 2020;370:m2516.

5. Deeks JJ, Dinnes J, Takwoingi Y, et al. Antibody tests for identification of current and past infection with SARS-CoV-2. Cochrane Database System Rev 2020;6:CD013652.

6. World Health Organization. Risk assessment and management of exposure of health care workers in the context of COVID-19: interim guidance, 19 March 2020. Geneva, Switzerland: World Health Organization, 2020. [https://apps.who.int/iris/handle/10665/331496]

7. Lahner E, Dilaghi E, Prestigiacomo C, et al. Prevalence of Sars-Cov-2 Infection in Health Workers (HWs) and Diagnostic Test Performance: The Experience of a Teaching Hospital in Central Italy. Int J Environ Res Public Health. 2020;17:4417.

8. Fill Malfertheiner S, Brandstetter S, Roth S, et al. Immune response to SARS-CoV-2 in health care workers following a COVID-19 outbreak: A prospective longitudinal study. J Clin Virol 2020 (in press).

9. Vandercam G, Simon A, Scohy A, et al. Clinical characteristics and humoral immune response in Healthcare Workers with COVID-19 in a Teaching Hospital in Belgium. J Hosp Infect 2020 Sept 11 (in press).

10. Jespersen S, Mikkelsen S, Greve T, et al. SARS-CoV-2 seroprevalence survey among 17,971 healthcare and administrative personnel at hospitals, pre-hospital services, and specialist practitioners in the Central Denmark Region. Clin Infect Dis 2020 (in press).

11. Fuereder T, Berghoff AS, Heller G, et al. SARS-CoV-2 seroprevalence in oncology healthcare professionals and patients with cancer at a tertiary care centre during the COVID-19 pandemic. ESMO Open 2020;5:e000889.

12. Fafi-Kremer S, Bruel T, Madec Y, et al. Serologic responses to SARS-CoV-2 infection among hospital staff with mild disease in eastern France. EBioMedicine 2020;59:102915.

13. Huang C, Wang Y, Li X, et al. Clinical features of patients infected with 2019 novel coronavirus in Wuhan, China. Lancet 2020;395:497–506.

14. Jiang F, Deng L, Zhang L, Cai Y, Cheung CW, Xia Z. Review of the Clinical Characteristics of Coronavirus Disease 2019 (COVID-19). J Gen Intern Med 2020;35:1545–9.

15. Meng X, Deng Y, Dai Z, Meng Z. COVID-19 and anosmia: A review based on up-to-date knowledge. Am J Otolaryngol 2020;41:102581.

16. Sun Y, Koh V, Marimuthu K, Ng OT, Young B, et al. Epidemiological and Clinical Predictors of COVID-19. Clin Infect Dis 2020;71:786–92.

17. Lan FY, Filler R, Mathew S, et al. COVID-19 symptoms predictive of healthcare workers’ SARS-CoV-2 PCR results. PLoS One 2020;15:e0235460.

18. Boffetta P, Violante F, Durando P, et al. Determinants of SARS-CoV-2 infection in Italian healthcare workers: a multicenter study. Submitted for publication.

19. http://opendatadpc.maps.arcgis.com/apps/opsdashboard/index.html#/b0c68bce2cce478eaac82fe38d4138b1

20. Watson J, Whiting PF, Brush JE. Interpreting a Covid-19 test result. BMJ 2020;369:m1808.

